# Evaluation of the General Practice Pharmacist (GPP) intervention to optimise prescribing in Irish primary care: a non-randomised pilot study

**DOI:** 10.1101/19009910

**Authors:** Karen Cardwell, Susan M Smith, Barbara Clyne, Laura McCullagh, Emma Wallace, Ciara Kirke, Tom Fahey, Frank Moriarty, on behalf of the General Practice Pharmacist (GPP) Study Group

**Affiliations:** HRB Centre for Primary Care Research, Department of General Practice, Royal College of Surgeons in Ireland, Dublin, Ireland; HRB Collaboration in Ireland for Clinical Effectiveness Reviews (HRB-CICER), Health Information and Quality Authority, Dublin, Ireland; National Centre for Pharmacoeconomics, St James’s University Teaching Hospital, Dublin, Ireland; Department of Pharmacology and Therapeutics, Trinity Centre for Health Sciences, Trinity College Dublin, Dublin, Ireland; National Quality Improvement Team, Health Service Executive, Dr. Steevens Hospital, Dublin

**Keywords:** Primary care, Organisation of health services, Quality in health care, Health economics

## Abstract

**Objective:** Limited evidence suggests integration of pharmacists into the general practice team could improve medicines management for patients, particularly those with multimorbidity and polypharmacy. This study aimed to develop and assess the feasibility of an intervention involving pharmacists, working within general practices, to optimise prescribing in Ireland.

**Design:** Non-randomised pilot study

**Setting:** Primary care in Ireland

**Participants:** Four general practices, purposively sampled and recruited to reflect a range of practice sizes and demographic profiles.

**Intervention:** A pharmacist joined the practice team for six months (10 hours/week) and undertook medication reviews (face-to-face or chart-based) for adult patients, provided prescribing advice, supported clinical audits, and facilitated practice-based education.

**Outcome measures:** Anonymised practice-level medication (e.g. medication changes) and cost data were collected. Patient-Reported Outcome Measure (PROM) data were collected on a subset of older adults (aged ≥65 years) with polypharmacy using patient questionnaires, before and six weeks after medication review by the pharmacist.

**Results:** Across four practices, 787 patients were identified as having 1,521 prescribing issues by the pharmacists. Issues relating to potentially inappropriate or high-risk prescribing were addressed most often by the prescriber (51.8%), compared to cost-related issues (7.5%). Medication changes made during the study equated to approximately €57,000 in cost savings assuming they persisted for 12 months. Ninety-six patients aged ≥65 years with polypharmacy were recruited from the four practices for PROM data collection and 64 (66.7%) were followed up. There were no changes in patients’ treatment burden or attitudes to deprescribing following medication review, and there were conflicting changes in patients’ self-reported quality of life.

**Conclusions:** This non-randomised pilot study demonstrated that an intervention involving pharmacists, working within general practices is feasible to implement and has potential to improve prescribing quality. This study provides rationale to conduct a randomised controlled trial to evaluate the clinical and cost-effectiveness of this intervention.

**Article summary:** 

**Strengths and limitations of this study:**

- This is the first study examining the role of general practice-based pharmacists in Ireland and the feasibility of evaluating this role.
- Integration of pharmacists was limited to four general practices, although these were diverse in terms of practice characteristics.
- A range of medication and patient-reported outcome measures data were collected, although because this was a pilot study there was no control group to compare these to.

## Introduction

The global burden of chronic disease is increasing, with more individuals living with long-term chronic conditions. Managing patients with multiple conditions (multimorbidity) and polypharmacy is recognised as a major challenge for healthcare systems.[1] Within general practice, challenges include the fragmentation of healthcare across the primary-secondary care interface, single-disease clinical guidelines which do not reflect multimorbid patients, challenges in delivering patient-centred care and shared decision-making.[2] General Practitioners (GPs) may also be co-ordinating prescribing from multiple specialists and attempting to balance benefits and risks from several medications. Thus, there has been an increased emphasis on the need to support GPs in the management of these patients. Appropriate medication use is one area of particular importance,[3] and polypharmacy has been identified as a major priority by the World Health Organisation through its third global patient safety challenge, Medication Without Harm.[4,5]

Interventions to improve appropriate prescribing include those aimed at GPs (e.g. computerised decision support),[6,7] patient educational interventions and changes to care delivery arrangements, such as staffing models or skills-mix.[8] Pharmacists, integrated within general practice as a member of the primary care team, may support appropriate medicines use and provide benefits to patients.[9,10] General practice-based pharmacists exist in a number of health systems internationally, including Canada, the United Kingdom (UK), and the United States of America (USA). Evidence from published evaluations suggests pharmacists in general practice can have a positive impact on clinical outcomes, such as blood pressure and glycosylated haemoglobin,[11] and may reduce medication-related hospitalisations.[12] They may also release GP capacity by reducing prescribing activities.[13] The model of practice-based pharmacists may provide advantages over a community pharmacy service, including (co-)location, access to medical records to inform the quality and appropriateness of recommendations, the potential for formal and informal communication and discussion of the pharmacist’s recommendations, and reduced fragmentation of care.[10,14–16]

Unlike countries such as the UK, pharmacists in Ireland have not been formally integrated into the general practice team, nor do they have prescribing rights. This feasibility study is part of a programme of work that follows the Medical Research Council (MRC) guidelines on developing and evaluating a complex intervention. The ultimate aim is to conduct a definitive randomised controlled trial (RCT) to compare the effectiveness and cost effectiveness of a General Practice Pharmacist compared to usual GP care. Therefore, this study aims to assess the feasibility and potential cost and clinical effectiveness of pharmacists, working with GPs, to optimise prescribing in Irish primary care.[17]

## Methods

### Study design and setting

This non-randomised pilot study was conducted following the principles of the Consolidated Standards of Reporting Trials (CONSORT) guidelines extension for the reporting of randomised pilot and feasibility studies.[18] Ethical approval was granted by the Irish College of General Practitioners and the study protocol has been published previously.[17]

This study was conducted in general practice in Ireland, which has a mixed private and public healthcare system. The state provides some individuals with access to medical services, including hospital inpatient and outpatient care, GPs and dental services, free at the point of use through the General Medical Services (GMS) scheme. Approximately 40% of the population is eligible based on household income, with a higher threshold applying to those aged ≥70 years so a greater proportion of this age group are covered. For people with income levels just above the GMS scheme threshold, a Doctor Visit Card (DVC) is granted which entitles them to only GP visits free of charge. The Health Service Executive (HSE) is the main provider of health services, particularly in secondary care, while in primary care, GPs and community pharmacists are private contractors who provide services for the HSE. The remainder of the population not covered by state schemes pay out-of-pocket for their care or may have voluntary private health insurance which can cover some or all costs.

### Participants

The study was conducted in four general practices. Practices were purposively selected from the national Primary Care Clinical Trials Network Ireland, to reflect a range of practice sizes and types, from both socioeconomically deprived and affluent areas. Practices were invited to participate by email (which included a study information sheet, practice consent form and practice profile questionnaire) and a follow-up telephone call from the principal investigator (SS). Consenting practices were visited by the study manager (KC) to discuss the logistics of the study and answer any further questions relating to their involvement in the study. Practice-related costs of participation in the study (room rental and GP time) were covered.

### Intervention components

Following the enrolment of practices in the study, a pharmacist joined the general practice team for a period of six months working ten hours per week, between September 2017 and March 2018. Pharmacists were recruited by the research team through an open recruitment process. The configuration of this time and activities in the practice were agreed between each practice and pharmacist. Unlike other countries, there are no formal training pathways or programs for practice-based pharmacists in Ireland. However each pharmacist was provided with a Study Intervention Manual which detailed the scope of activities to be delivered by the pharmacist, based on national guidance and previous research.[19–21] Broadly, this involved medications reviews (both opportunistic and targeted, conducted face-to-face with patients or using patients’ medical charts only), involvement in the repeat prescribing process, conducting educational sessions with general practice staff and supporting GPs in undertaking clinical audits.

It was recommended that the medication reviews focussed on three domains, (i) high-risk prescribing practices [22] and potentially inappropriate prescribing (PIP),[17,23] (ii) deprescribing of medications that may cause harm or are no longer providing benefit,[24] and (iii) rational and cost-effective prescribing, including use of ‘preferred drugs’ in accordance with recommendations from the HSE Medicine Management Programme (MMP).[19,20] The Preferred Drug Initiative has identified a single ‘preferred drug’ within several therapeutic drug classes, based on clinical- and cost-effectiveness, which prescribers are recommended to use where possible, e.g. lansoprazole as the preferred proton pump inhibitor. The MMP also provides recommendations on which dose-equivalent inhalers for obstructive airway conditions are most cost-effective. Pharmacists used indicators pre-specified in the study intervention manual to screen the medical records of the practice patient population and identify potential issues. Pharmacists do not have prescribing rights in Ireland, and therefore the GP maintained clinical autonomy and implemented any changes to prescriptions, typically with non-urgent issues addressed at patients’ next appointments and patients being contacted where more immediate changes were required.

### Outcome data collection

#### Prescribing data

Throughout the six-month study intervention period, pharmacists collected demographic and medication data for various prescribing indicators (as defined in the Study Intervention Manual) relating to adults patients of any age. These prescribing indicators were related to (i) prescribing practices that were considered potentially inappropriate (i.e. PIP) or high risk, (ii) instances where medicines could be appropriately deprescribed and (iii) instances where Preferred Drugs could be prescribed instead of non-preferred drugs. These data were collected whilst undertaking chart-based and face-to-face medication reviews, were anonymised with no patient identifiers, and were recorded in a pre-defined data collection sheet. eTables 1, 2 and 3 describe the prescribing indicators used in this study.

#### Patient-reported outcome measure (PROM) data

Patient-reported outcome measure (PROM) data were collected in addition to the prescribing data from medication reviews to assess the feasibility of such data collection in a future randomised trial. In month 4 of the study period, eligible patients were invited to have a medication review conducted by the pharmacist, and were asked to complete a patient questionnaire before and three months after this review.

Using the practices’ prescribing software, pharmacists compiled a list of patients aged ≥65 years who were taking ≥10 repeat medicines. Thereafter, these patients were screened by the pharmacist and a GP to ensure they were able to provide informed consent and participate in data collection. Patients were excluded if they had psychiatric or psychological morbidity or cognitive impairment sufficient to impair the provision of informed consent, a terminal illness likely to lead to death or major disability during the study follow-up period, or if they had already been reviewed by the pharmacist. Eligible patients were invited to participate via a letter (sent from the general practice) and received a follow-up call from a member of the practice administration staff. Following the provision of informed consent, patients were scheduled for a face-to-face medication review with the pharmacist. Chart-based medication reviews were completed for those patients who declined to attend for a face-to-face medication review. Chart-based reviews and lowering the repeat medicines threshold to ≥7, and then ≥5 medications, were protocol amendments implemented due to poor response rates.

The baseline patient questionnaire included questions on demographics, healthcare utilisation, and PROMs relating to quality of life (EQ-5D-5L and visual analogue scale [VAS]) and medications (the Multimorbidity Treatment Burden Questionnaire [MTBQ] and the revised Patient Attitudes Towards Deprescribing [rPATD]).[25–27] The MTBQ is a 13-item questionnaire which measures treatment burden (the effort of looking after one’s health) in patients with multimorbidity in primary care, while the rPATD questionnaire contains 22 items to capture older adults’ beliefs and attitudes towards deprescribing. Only the PROMs were included in the follow-up questionnaire at six weeks post review.

### Data analysis

Descriptive statistics for general practices and patients with a prescribing issue were generated. For each prescribing indicator we summarised the prevalence as a percentage of all the indicators within that category of prescribing issue. The proportion of cases where the implicated medication had been prescribed long-term (i.e. prescribed for ≥6 months) and where the prescribing issue was addressed during the study period were also determined (i.e. where a GP made some change to the medications following the pharmacist’s intervention). Any changes occurring after the end of the six month study period were not captured. For PROM data collection, differences in outcomes reported pre- and post-intervention were summarised. For the EQ-5D-5L, participants were classed into those whose health state improved (improvement in at least one dimension and no worse in any other), worsened (worsening in at least one dimension and no better in any other), no change, or mixed change (improved and worsened in different health states).[28] MTBQ was summarised using median and interquartile range, and by classifying into burden categories (none, low, medium and high), as recommended by the tool’s developers due to non-normality.[26] Changes in EQ-VAS, EQ-5D-5L utility score (based on an Irish value set derived from a general population representative sample),[29] and each dimension of the rPATD were examined using a paired-samples t-test. Data analysis was conducted using Stata version 14 and p-values <0.05 were deemed significant.

A cost analysis was conducted to determine the costs saved or incurred for changes in prescribing due to the GPP intervention. This analysis was based on the cost of providing the intervention and the cost savings realised from the intervention. For each medication change detected during the study period, the total cost to the health system over twelve months was calculated. This cost included the publically-available drug reimbursement price (less wholesaler discount),[30] as well as pharmacist dispensing fees of €5 per item (in line with the HSE dispensing fee structure).[31] Where medications were amended, the cost difference between the original and new prescription was calculated. Costs of laboratory tests ordered as a result of pharmacist review were also determined. Where data on the original and/or new prescription were missing, the mean cost difference for changes within that indicator was used. This analysis only considered direct costs relating to medication changes and did not evaluate the downstream savings due to more appropriate prescribing, such as reduced medication-related hospitalisations.

### Continuation criteria

Continuation criteria were used to determine whether further evaluation of this intervention is justified. These continuation criteria were outlined in the study protocol paper,[32] based on consideration of the primary objectives around feasibility (including recruitment and retention of practices, pharmacists and patients) and the potential for effectiveness and system-wide implementation.

### Sample size

Since this was a feasibility study, no formal power calculation was conducted. The recruitment target for the PROM data collection was 50 participants per practice (200 in total), however, this target was not reached.

### Patient involvement

There was no formal public and patient involvement for this study, however we engaged with the Collaborative Doctoral Awards in Multimorbidity PPI panel for consideration of the continuation criteria and input into the follow-on pilot cluster RCT.

## Results

Three pharmacists were integrated into four participating general practices for a period of six months; one pharmacist delivered the intervention in two practices. Pharmacists (one male, two female) had a mean of 15.7 years’ clinical experience as a pharmacist prior to their involvement in the study. Recruited general practices were from both socioeconomically deprived and affluent areas; see Table 1 for an overview of the characteristics of enrolled practices at baseline.

**Table 1.**
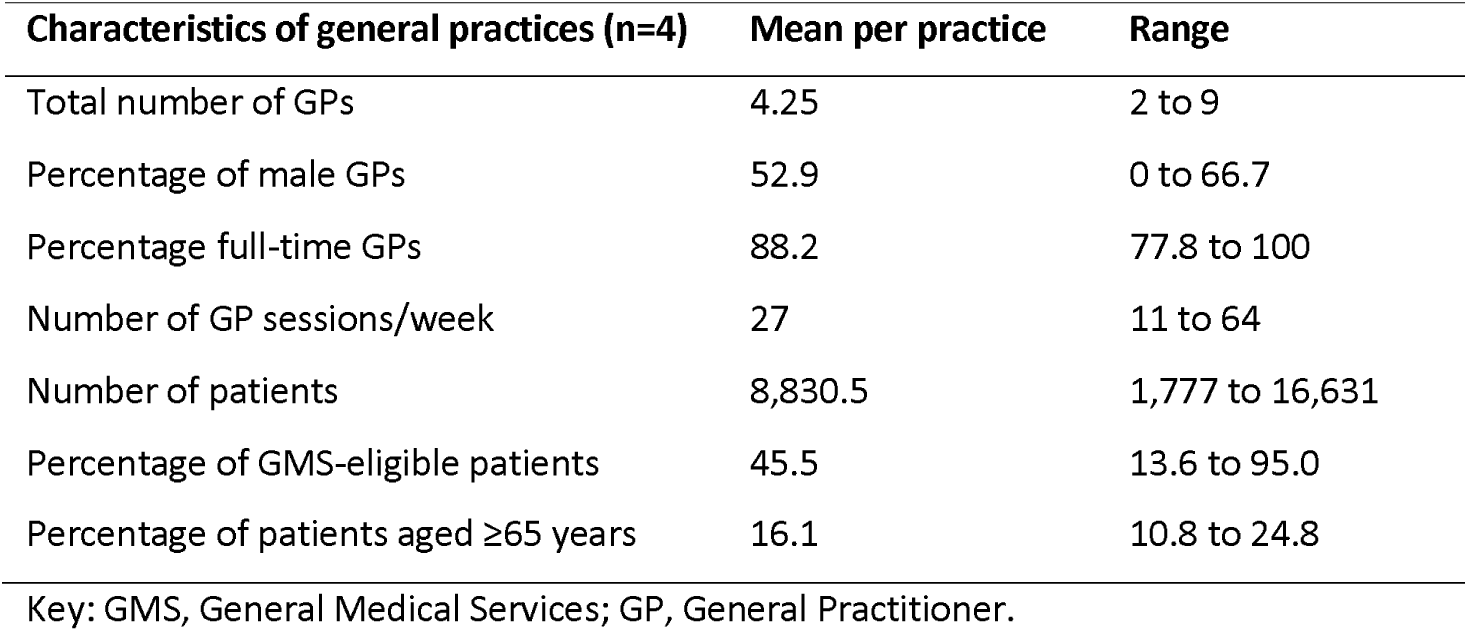
Baseline characteristics of the general practices enrolled in the study.

| Characteristics of general practices (n=4) | Mean per practice | Range |
| --- | --- | --- |
| Total number of GPs | 4.25 | 2 to 9 |
| Percentage of male GPs | 52.9 | 0 to 66.7 |
| Percentage full-time GPs | 88.2 | 77.8 to 100 |
| Number of GP sessions/week | 27 | 11 to 64 |
| Number of patients | 8,830.5 | 1,777 to 16,631 |
| Percentage of GMS-eligible patients | 45.5 | 13.6 to 95.0 |
| Percentage of patients aged $\geq 65$ years | 16.1 | 10.8 to 24.8 |

Key: GMS, General Medical Services; GP, General Practitioner.

In three practices, pharmacists were based in their own room with their own computer and had access to patients’ electronic medical records. In one practice, the pharmacist was based either in the practice administration office with the administration staff or in their own room, depending on the availability of practice rooms on that day. Tasks undertaken by the pharmacist at each practice included identifying potential prescribing issues (both those pre-specified in the Study Intervention Manual and others based on their clinical judgement) and facilitation of practice audits. In addition, one pharmacist delivered practice-based educational sessions on the treatment and management of patients with chronic obstructive pulmonary disease or type II diabetes mellitus, using an electronic prescribing tool developed by the pharmacist and a GP at that practice. One intervention component (management of repeat prescribing) was not delivered by any pharmacist at any recruited practice, as this process had been standardised and was operating successfully within each practice.

### Medication reviews

Pharmacists identified 786 patients with one or more prescribing issue during chart-based or face-to-face medication reviews, the majority via chart-based review (n=748). The mean age was 69.8 years (SD 14.8), and 65.2% were female (n=513). The majority (649, 82.4%) were GMS patients, while 42 (5.3%) were DVC patients. Patients were on a mean of 9.5 medications (SD 5.5), with a mean of 4.7 medications (SD 3.6) prescribed generically.

A total of 1,521 potential issues were identified (Table 2), a total of 59.6% relating to high-risk or potentially inappropriate prescribing, 9.5% where medicines could be deprescribed, and 31.0% where a non-preferred drug was prescribed. The most common PIP/high-risk prescribing indicators identified involved long-term proton pump inhibitors at maximal dose, short-acting benzodiazepines, non-steroidal anti-inflammatories, the prescribing of duplicate therapeutic classes in the same patient and tricyclic antidepressants. Most medications involved had been prescribed for >6 months (eTable 4). Among the most common prescribing indicators, duplicate therapeutic drug classes were most commonly addressed (within the study period) following the pharmacist’s intervention (in 87.9% of cases) while short-acting benzodiazepines were least frequently addressed (in 11.4% of cases). The most common deprescribing indicators identified were ‘Other’ (this category included several drugs that were no longer required by/indicated for the patient e.g. quinine sulphate, cyclizine, domperidone, ferrous fumarate and valsartan), z-drugs, antihistamines, betahistine and bisphosphonates. All medications involved had been prescribed for >6 months. In relation to medications that could be deprescribed, other drugs (most often calcium and vitamin D combinations), z-drugs, antihistamines, and betahistine were most commonly identified by the pharmacist. All of these were addressed by the prescriber through dose reduction or stopping in the majority of cases, with the exception of Z-drugs which were only addressed in 17.9% of cases. The most common drug categories in which non-preferred drugs were prescribed were statins, angiotensin II receptor blockers, proton pump inhibitors, selective serotonin reuptake inhibitors and beta-blockers. Among all the indicators identified, only non-preferred inhalers and proton pump inhibitors were addressed (in 75.0% and 24.0% of cases, respectively).

**Table 2.**
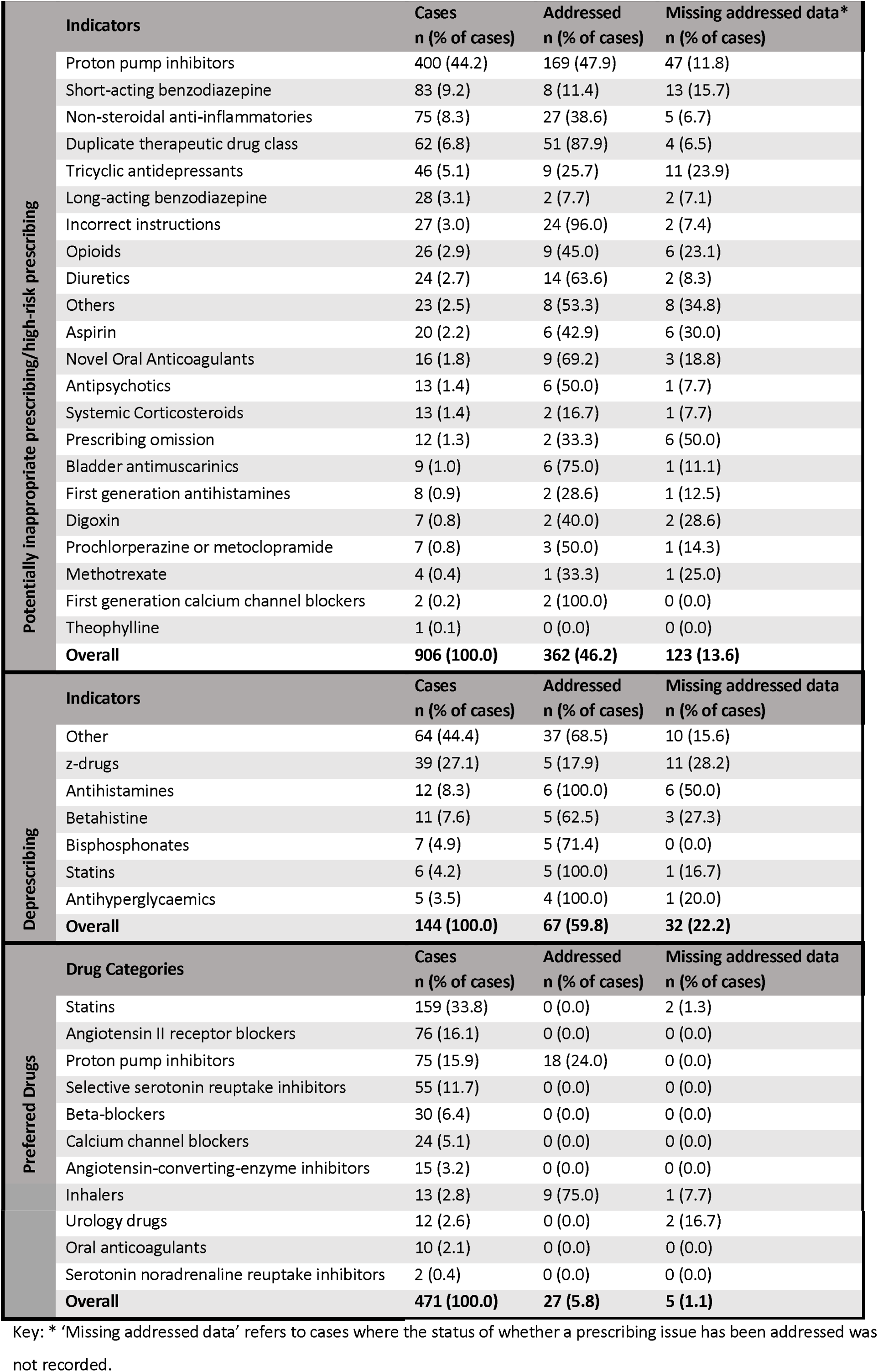
Prescribing issues identified by pharmacists during medication reviews (both chart-based and face-to-face).

### Patient reported outcome measures

Ninety-six patients of a pre-specified target of 200 (48%) were reviewed as part of PROM data collection. Table 3 describes the characteristics of these patients and their detailed self-reported healthcare utilisation over the 12 months prior to the study is included in the Supplementary Material. Sixty-four patients completed a questionnaire six weeks following their medication review. Table 4 compares patients’ quality of life and attitudes towards medicines pre- and post-review, while the distribution across dimensions and levels of EQ-5D-5L is show in eTable 5. There was a statistically significant reduction of 0.06 in EQ-5D-5L score (95% confidence interval -0.11 to - 0.002), however when measured using the EQ-VAS, quality of life increased by a similar magnitude (0.06, 95% CI 0.02 to 0.10). Overall, there was no significant change in Multimorbidity Treatment Burden Questionnaire score post-review compared to pre-review, and the distribution of patients across levels of burden did not change. Regarding attitudes towards deprescribing, patients scored the appropriateness and involvement factors higher than the burden and concerns factors, however neither these nor the two global items changed significantly following the intervention.

**Table 3.** Characteristics of patients enrolled in the PROM study and who completed patient questionnaires.

| **Characteristic** | **n (%)** |
| --- | --- |
| **Age, mean (SD)** | 77.7 (6.4) |
| **Female gender** | 63 (65.6) |
| **Marital status** |  |
| Married or cohabiting | 48 (50.0) |
| Widowed, divorced or separated | 32 (33.3) |
| Single/never married | 17 (17.7) |
| **Irish nationality** | 90 (93.8) |
| **Education** |  |
| No schooling or primary only | 31 (32.3) |
| Secondary level | 32 (33.3) |
| Third level | 33 (34.4) |
| **Employment** |  |
| Retired | 71 (74.0) |
| Employed or homemaker | 21 (21.9) |
| Other | 4 (4.2) |
| **Private health insurance** | 46 (47.9) |
| **GMS scheme** | 65 (67.7) |
| **Doctor visit card** | 19 (19.8) |
| **Distance GP, median (IQR)** | 2 (1 to 4) |
| **Any GP visit in previous 12 months** | 95 (99.0) |
| Number of visits, median (IQR) | 5.5 (3.5, 8) |
| **Any A&E visit in previous 12 months** | 27 (28.1) |
| Number of visits, median (IQR) | 1 (1, 2) |
| **Any outpatient visit in previous 12 months** | 52 (54.2) |
| Number of visits, median (IQR) | 2 (1, 6) |
| **Any inpatient stay in previous 12 months** | 29 (30.2) |
| Number of stays, median (IQR) | 1 (1, 3) |
| Number of inpatient nights, median (IQR) | 3 (1, 8) |

Key: A&E, accident & emergency department; GMS, General Medical Services; SD, Standard deviation; IQR, Interquartile range.

**Table 4.** Comparison of patients’ perceived level of health and attitudes towards their medicines pre- and post- medication review with the pharmacist.

| Patient-reported outcome measure | n | Pre-review | Post-review | Mean difference (95% CI) |
| --- | --- | --- | --- | --- |
| **EQ-5D-5L** |  |  |  |  |
| Utility score<sup>a</sup>, mean | 62 | 0.728 | 0.673 | -0.056 (-0.111 to -0.002)* |
| Health states change, n (%) | 64 |  |  |  |
| No change | 8 (12.5) |  |  |  |
| Improve | 19 (29.7) |  |  |  |
| Worsen | 23 (35.9) |  |  |  |
| Mixed change | 14 (21.9) |  |  |  |
| **EQ-VAS, mean** | 60 | 0.641 | 0.701 | 0.061 (0.018 to 0.103)* |
| **MTBQ** |  |  |  |  |
| Score, median (IQR) | 64 | 5.8 (0 to 11.5) | 5.8 (1.9 to 16.3) |  |
| Burden categories, n (%) | 64 |  |  |  |
| No burden |  | 17 (26.6) | 17 (26.6) |  |
| Low burden |  | 30 (46.9) | 26 (40.6) |  |
| Medium burden |  | 11 (17.2) | 11 (17.2) |  |
| High burden |  | 6 (9.4) | 10 (15.6) |  |
| **rPATD, mean** |  |  |  |  |
| Burden | 54 | 13.03 | 13.31 | 0.28 (-0.71 to 1.26) |
| Appropriateness | 52 | 17.13 | 17.23 | 0.10 (-0.73 to 0.92) |
| Concerns | 54 | 12.00 | 12.33 | 0.33 (-0.95 to 1.61) |
| Involvement | 60 | 21.33 | 21.6 | 0.27 (-0.62 to 1.15) |
| Satisfaction | 62 | 4.13 | 4.11 | -0.02 (-0.29 to 0.25) |
| Willingness | 63 | 4.54 | 4.37 | -0.17 (-0.36 to 0.01) |

Key: EQ-5D-5L, EuroQol-Health-related Quality of life five level questionnaire; EQ-VAS, EuroQol-visual analogue scale; MTBQ, Multimorbidity Treatment Burden Questionnaire; rPATD, Revised Patients' Attitudes Towards Deprescribing, CI, Confidence Intervals. <sup>a</sup> Using an Irish value set derived from a general population representative sample.[29] \* paired t-test <0.05

### Cost data

The total cost of three pharmacists’ salaries across four practices was €31,200. This was based on €30/hour for 10 hours/week over 26 weeks. Practice-related costs for study participation included room rental (€15/hour for 10 hours/week) and GP time for meeting with the pharmacist (€70/hour for 1 hour/week), amounting to €22,880. Table 5 reports the cost-savings realised from addressed prescribing interventions. Overall cost-savings that would accrue over a 12-month period following prescription changes amounted to €56,669.

**Table 5.** Cost-savings (over a 12-month period) from prescribing interventions

| Cost category | Cost saving | Cost incurred |
| --- | --- | --- |
| Cost savings for amended drugs* | €6,374 |  |
| Cost savings for stopped drugs | €43,681 |  |
| Cost savings from interventions lacking full information | €7,162 |  |
| **Total cost savings for drug intervention** | **€57,217** |  |
| Cost of tests ordered |  | €549 |
| **Overall savings from intervention** | **€56,669** |  |

Key: \* Cost of amended drugs was €31,394 before intervention and €25,019 after intervention.

### Continuation criteria

All continuation criteria, with the exception of two, reached the threshold of ‘Proceed with RCT’, including those relating to practice and pharmacist recruitment and retention, and intervention acceptability, feasibility, and potential for cost savings (see supplementary files, eFigure 1). Those relating to patient recruitment and retention for PROM data collection were at the level of ‘Do not proceed with RCT unless problems can be overcome’.

## Discussion

### Summary of findings and context in the literature

To our knowledge this feasibility study is the first to introduce and evaluate the feasibility of pharmacists in the general practice setting to support prescribing in Ireland. A large number of prescribing issues were identified by the pharmacists, however the extent to which these were addressed by the prescriber differed depending on the nature of the issues, ranging from 7.5% of those relating to cost-effectiveness (i.e. use of non-preferred drugs within a class) to 51.8% of those relating to potentially inappropriate or high-risk prescribing. The medication changes detected during the study period equated to approximately €57,000 in cost savings assuming they persisted for 12 months. Although this feasibility study was not adequately powered to detect statistically significant outcome changes, these findings are encouraging and warrant further investigation to test the effectiveness of the intervention in future randomised studies.

Two recent systematic reviews found mixed evidence regarding the benefits of pharmacists in general practice to support prescribing, which may depend on heterogeneity in patient population included (i.e. those with specific medical conditions, or generally at risk of medication issues), outcomes assessed (i.e. clinical, surrogate, or patient-reported outcomes), and to what extent pharmacists were integrated into the general practice setting.[10,11] As highlighted in our study, the extent to which issues were addressed by prescribers differed, with issues of safety (i.e. potentially inappropriate or high-risk prescribing) addressed more than issues of cost.

There were low levels of changes in cases of non-preferred drugs (based on effectiveness, safety and cost-effectiveness) within a drug class being prescribed in our study. Non-preferred inhalers were changed in more than two thirds of cases, however this category was somewhat different, as it represented a change of formulation rather than other categories which involved a change of chemical entity, and was also the subject of an audit in one practice. Reviews on barriers and facilitators of deprescribing may offer some explanations for the relatively low uptake of these medication changes.[33,34] Evidence that it is often only after a medication problem has occurred that deprescribing is considered may suggest that cost savings to the health system is an insufficient motivator to switch medications for a patient who is a prevalent user if no issue has arisen.[33] Similarly, fear of negative consequences of a change (i.e. potential adverse reactions to a new agent) is another potential barrier.[34] By contrast, the safety concerns in cases of high-risk prescribing may have been sufficient to outweigh these fears resulting in higher uptake of these recommended changes.

There was a low response from patients to face-to-face review invitations with pharmacists as part of the PROM data collection with a recruitment rate of 48%. While qualitative evidence from England indicates that pharmacist-led polypharmacy medication reviews are a positive experience for older individuals,[35] the patients in this current study likely already had high treatment burden and frequent routine visits to healthcare professionals, given that they were on a high number of medications.[36] They therefore may have been reluctant to attend an additional appointment for the purpose of pharmacist review as part of this study. In addition, the burden of data collection as part of this study and unfamiliarity with the role of pharmacists in general practice may have hampered participation. The continuation criteria indicated the problems of patient recruitment and retention need to be overcome in order to proceed with further evaluation of the intervention. This will be addressed by recruiting patients at study commencement before a pharmacist is integrated into each practice, to ensure an adequate sample is achieved and to facilitate alignment of medication review with other routine visits.

There was a significant improvement in quality of life as measured by the EQ-VAS following the medication review and examining changes across EQ-5D-5L health states indicated no prevailing pattern of change following our intervention. This was inconsistent with the index score from applying an Irish value set to the EQ-5D-5L responses, however the use of an external value set introduces an external source of variance and the values and weighting of different dimensions may not reflect those of participants in this study.[28] The EQ-VAS captures health more broadly than the dimensions included in the EQ-5D-5L,[28] and has diverged from EQ-5D-5L index score in a previous trial of medication reviews in older adults.[37]

### Strengths and limitations

To our knowledge, this is the first study examining the role of practice-based pharmacists in Ireland and the feasibility of evaluating this role. We used a range of valid indicators of high-risk and potentially inappropriate prescribing to guide pharmacists’ interventions and to measure the potential clinical benefits. This pilot study was limited by its small sample size and pre-post design in relation to patient-reported outcomes without inclusion of a control group, however this was appropriate given the aim to assess feasibility. In addition, the generalisability of this study is limited by inclusion of only four purposively selected general practices. While uptake of pharmacist recommendations in relation to preferred drugs was low, a limitation of this study is that analysis only focussed on prevalent users of relevant drug classes, and therefore potential influence of pharmacists on prescribing in cases of new initiations could not be captured. The cost data captured was in the context of this research study, and did not account for real-world employment costs that would be associated with the introduction of such a role.

## Conclusions

This study found that the integration of pharmacists, working with GPs, to optimise prescribing in Irish primary care is largely feasible and has potential clinical and cost benefits. A qualitative evaluation of this feasibility study is ongoing to explore this role further and inform future research. In line with the MRC guidelines on developing and evaluating complex interventions, this will now proceed to a randomised pilot study, with changes to the intervention and study design informed by the results of the present study (in particular, relating to patient recruitment) and qualitative evaluation. This will provide further evidence on the role of pharmacist in GP practices in the Irish context, and the potential of this intervention to help achieve the ambitious target of the WHO’s Medication Without Harm challenge to reduce serious, avoidable medication-related harm by 50% in 5 years.[4]

## Data Availability

Data and analytical code relating to prescribing issues is available from www.doi.org/10.5281/zenodo.3492198.

## Acknowledgements

The other members of the GPP Study Group are Michael Barry, Fiona Boland, Sarah Clarke, Karen Finnigan, Maria Daly, Catriona Bradley, Paul Gallagher, Edel Murphy, Andrew Murphy, Patrick Byrne, Aisling Croke and Oscar James.

## Author contributions

SMS conceived the study and KC, SMS, BC, LMcC, EW, CK, TF, and FM contributed to the design of the study. KC coordinated the intervention delivery and data collection. KC, LMcC, and FM, analysed the data and all authors were involved in the interpretation of the data. The manuscript was drafted by FM and all authors were involved in the critical revision and approval of the final manuscript.

## Supplementary materials

eFigure 1. Continuation criteria results from feasibility study

eTable 1. Prescribing indicators that were defined as potentially inappropriate or high-risk in the General Practice Pharmacist Study

eTable 2. Deprescribing indicators used to identify drugs that could be considered for deprescribing in the General Practice Pharmacist Study.

eTable 3. Preferred drugs to prescribe as defined by the Health Service Executive Medicines Management Programme.

eTable 4 Prevalence and duration of prescribing issues identified by pharmacists during medication reviews (both chart-based and face-to-face).

eTable 5. Distribution of participants across EQ-5D-5L dimensions and levels, pre and post medication review.

